# How do local-level authorities engage in epidemic and pandemic preparedness activities and coordinate with higher levels of government? Survey results from 33 cities

**DOI:** 10.1101/2022.05.25.22275614

**Authors:** Matthew R Boyce, Melissa Cordoba Asprilla, Breanna van Loenen, Amanda McClelland, Ariella Rojhani

## Abstract

The COVID-19 pandemic suggests that there are opportunities to improve preparedness for infectious disease outbreaks. While much attention has been given to understanding national-level preparedness, relatively little attention has been given to understanding preparedness at the local-level. We, therefore, aim to describe (1) how local governments were engaged in epidemic preparedness efforts before the COVID-19 pandemic and (2) how they were coordinating with authorities at higher levels of governance before COVID-19. We developed an online survey and distributed it to 50 cities around the world involved in the Partnership for Healthy Cities. The survey included several question formats including free-response, matrices, and multiple-choice questions. RACI matrices, a project management tool that helps explain coordination structures, were used to understand the level of government responsible, accountable, consulted, and informed regarding select preparedness activities. We used descriptive statistics to summarize local-level engagement in epidemic preparedness. Local health authorities from 33 cities completed the survey. Prior to the COVID-19 pandemic, 20 of the cities had completed infectious disease risk assessments, 10 completed all-hazards risk assessments, 11 completed simulation exercises, 10 completed after-action reviews, 19 developed preparedness and response plans, three reported involvement in their country’s Joint External Evaluation of the International Health Regulations, and eight cities reported involvement in the development of their countries’ National Action Plan for Health Security. RACI matrices revealed various models of epidemic preparedness, with responsibility often shared across levels, and national governments accountable for the most activities, compared to other governance levels. In conclusion, national governments maintain the largest role in epidemic and pandemic preparedness but the role of subnational and local governments is not negligible. Local-level actors engage in a variety of preparedness activities and future efforts should strive to better include these actors in preparedness as a means of bolstering local, national, and global health security.

## Introduction

Ensuring that urban settings are prepared for public health emergencies, such as infectious disease outbreaks, is critically important. This is because, in addition to potentially being where outbreaks originate, characteristics of urban environments can promote the spread of disease, both within the city and globally [1-3]. In today’s interconnected world, if urban environments are not well capacitated and prepared to respond to such events, localized outbreaks can rapidly escalate into those of a much larger scale, such as epidemics or pandemics. The rapid, global spread of the SARS-CoV-2 virus and the ensuing COVID-19 pandemic do well to demonstrate this reality, as do other notable outbreaks including the 2003 SARS epidemic, the 2009 H1N1 influenza pandemic, and the 2015-2016 Zika epidemic [3,4]. Thus, public health preparedness in cities and urban environments is essential for ensuring local, national, and global health security.

While local authorities and public health departments in cities can play key roles in promoting preparedness and preventing the spread of infectious diseases, they are not the only entities involved in preparedness efforts. Much attention has been given to national-level public health preparedness and health security efforts, such as the International Health Regulations Monitoring and Evaluation Framework [5]. This Framework includes four core components: mandatory self-assessments, voluntary external assessments (i.e., the Joint External Evaluation (JEE)), voluntary after-action reviews, and voluntary simulation exercises. These components seek to improve preparedness by assessing the existence (i.e., self and external assessments) and functionality (i.e., after-action reviews and simulation exercises) of the capacities essential for detecting, assessing, notifying and responding to public health emergencies [6].

Before the COVID-19 pandemic, many countries reported high levels of preparedness through these assessments. Other assessments, such as the Global Health Security Index and the Epidemic Preparedness Index, also suggested that many countries were well prepared for an epidemic or pandemic, especially those in Europe and North America [7,8]. Still, initial results suggest that these measures of preparedness were not highly correlated with pandemic response performance [9-12]. Among the emerging hypotheses for why these preparedness assessments were not correlated with pandemic response performance is that they were too reductive and did not sufficiently account for specific and local-level contexts and inequalities [10,11]. The health, economic, and social consequences that have resulted from the pandemic further suggest that there are opportunities to improve public health preparedness [13,14].

In addition to overlooking local-level contexts in preparedness assessments, the roles of local authorities in these aforementioned efforts and the coordination between different levels of government remain poorly understood. This gap is notable given the widely recognized role that local authorities play in implementing policy decisions and producing the outcomes associated with governance processes, including the protection of public health [15]. In much of the world, public health is a local, community-based endeavor that depends on trust to effectively implement policies and deliver services [12]. For instance, many of the capacities measured in the preparedness assessments represent essential public health functions that are often carried out or supported by local authorities, such as disease surveillance, risk communication, and public health workforce development and maintenance [16,17].

The actual or potential actions that may be taken by local governments are often influenced by policy, guidance, and financing from higher levels of governance [18,19]. In many cities, total autonomy from higher levels of government is unlikely, and coordination between these actors is, therefore, an important consideration. This is especially true as inadequate coordination and poor governance can lead to implementation challenges, the inefficient use of limited resources, and greater health consequences [4,16,20,21].

These considerations lead to questions about not only how local governments and authorities were engaging in epidemic preparedness before the COVID-19 pandemic, but also the roles they played and how these efforts were coordinated with higher levels of government. The objectives of this descriptive research, then, are to understand the ways in which local-level authorities were engaging in preparedness efforts before the COVID-19 pandemic, as well as how they were coordinating with authorities at higher levels of governance to complete key tasks and preparedness activities.

## Methods

The research described in this article used a survey study design to ask local public health authorities in cities around the world about their city’s engagement in pandemic preparedness efforts prior to the COVID-19 pandemic and how select preparedness activities were coordinated with higher levels of government.

### Study Population

This research effort involved public health authorities from cities that were a part of the Partnership for Healthy Cities. The Partnership for Healthy Cities is a city network of 70 cities that have committed to preventing noncommunicable diseases and injuries through proven, evidence-based interventions and approaches. In March of 2020, recognizing the urgency of COVID-19 response, the network temporarily expanded its scope to include support for pandemic response. The population involved in this study included 50 cities in the Partnership for Healthy Cities that were purposively selected based on their involvement in a mini-grant program that focused on bolstering the local response to the COVID-19 pandemic.

### Questionnaire Development and Distribution

We developed a survey questionnaire based on existing pandemic preparedness frameworks and guidance [22,23]. The survey contained 17 questions – relating to participant demographics and preparedness efforts and activities – that included a variety of formats including free-response, matrices, and multiple-choice questions. The survey questionnaire is available for review in the S1 Appendix.

Included in these questions were RACI (i.e., Responsible, Accountable, Consulted, and Informed) matrices [24]. RACI matrices are project management tools that are useful for explaining coordination structures and the hierarchical nature of activities. Multiple levels of government can be responsible, consulted, and informed for a given activity. However, only one level of government can be accountable for a given activity. These queries allowed for survey responses to provide a granular look at how local preparedness activities related to and were coordinated with those of higher levels of government.

For the purposes of this study, we defined responsibility as implementing the work required to complete an activity; accountability was defined as overseeing the correct and thorough completion of an activity; consulted was defined as engaging in two-way communication to provide information necessary for the completion of an activity; and informed was defined as being updated on progress toward or on the results from a given activity (i.e., one-way communication) [24]. Additionally, we defined the national-level as relating to the efforts of the national government; the subnational-level relating to the efforts of state, provincial, district, or county governments; and the local-level relating to the efforts of city or municipal governments.

Once finalized, we translated the questionnaire and accompanying instructions into French and Spanish in an effort to ensure that the materials were accessible and comprehensible to potential survey participants.

### Ethics Statement

We submitted all study materials to the Georgetown University Institutional Review Board for ethical review. The Institutional Review Board granted an exemption to the study (STUDY00003948) in June 2021 after determining that the research posed no more than minimal risk to those who would be participating. As the study was granted exemption, informed consent was not required to be obtained. However, as a best practice in research, we provided an informed consent form to all study participants that emphasized the voluntary nature of their participation and that they maintained the option of withdrawing from the study at any time without consequence. Consent was then provided through participation.

### Data Collection & Analysis

We uploaded the survey to Qualtrics (Seattle, WA) and distributed it with instructions to local-level authorities, before closing the survey in August 2021. We then collected the data from survey responses and compiled them in a spreadsheet created with Microsoft Excel (Redmond, WA). Prior to beginning data analysis, we reviewed data for consistency and validity. We used Microsoft Excel and STATA v.17BE (College Station, TX) to perform basic descriptive statistical analyses to summarize local-level engagement in preparedness efforts and how they coordinated with higher levels of government to complete preparedness activities.

## Results

Local-level health authorities from 33 cities – Abidjan, Accra, Addis Ababa, Amman, Athens, Bandung, Bangkok, Barcelona, Bengaluru, Buenos Aires, Cali, Chicago, Colombo, Fortaleza, Guadalajara, Harare, Kampala, Kigali, Kumasi, León, Lima, London, Lusaka, Melbourne, Medellín, Montevideo, Ouagadougou, Quezon City, Rio de Janeiro, Santiago, Santo Domingo, Vancouver, and Yangon – completed surveys. Eight (24.2%) of these cities are located in countries that are classified as high-income countries, 12 (36.4%) are located in countries that are classified as upper-middle income countries, nine (27.3%) are located in countries that are classified as lower-middle income countries, and four (12.1%) are located in countries that are classified as low income countries (Table 1). A majority of survey participants were female and had worked in their current professional role for 1 to 4 years.

**Table 1.** Characteristics of survey participants and cities (n = 33).

| Characteristic | n (%) |
| --- | --- |
| Gender of Survey Participant |  |
| *Male* | 13 (39.4) |
| *Female* | 20 (60.6) |
| *Other/Prefer not to say* | 0 (0) |
| Number of Years Served in Current Role |  |
| *Less than 1 year* | 7 (21.2) |
| *1 to 4 years* | 16 (48.5) |
| *5 to 9 years* | 7 (21.2) |
| *10 years or more* | 3 (9.1) |
| Economy Income Level |  |
| *High income* | 8 (24.2) |
| *Upper-middle income* | 12 (36.4) |
| *Lower-middle income* | 9 (27.3) |
| *Low income* | 4 (12.1) |

### Local-Level Epidemic and Pandemic Preparedness Efforts

The preparedness experiences of cities varied across the world, reflecting the heterogeneity in the geographic, epidemiologic, political, and development contexts in which the cities exist. No city reported that they had completed or participated in all seven of the preparedness efforts. The greatest number of preparedness efforts reported was five of the seven, which was reported by Chicago, Colombo, and Melbourne.

Twenty of the cities (60.6%) reported that they had completed an infectious disease risk assessment prior to the COVID-19 pandemic, 10 (30.3%) reported the completion of an all-hazards risk assessment prior to the COVID-19 pandemic, 11 (33.3%) reported the completion of a simulation or table-top exercise, and 10 (30.3%) reported the completion of an after-action review prior to the COVID-19 pandemic (Fig 1). Nineteen (57.6%) of the cities also reported that they had developed a city-specific preparedness and response plan before the COVID-19 pandemic, often for other infectious diseases that represent epidemic or pandemic threats, such as cholera, influenza, or Zika.

**Fig 1.**
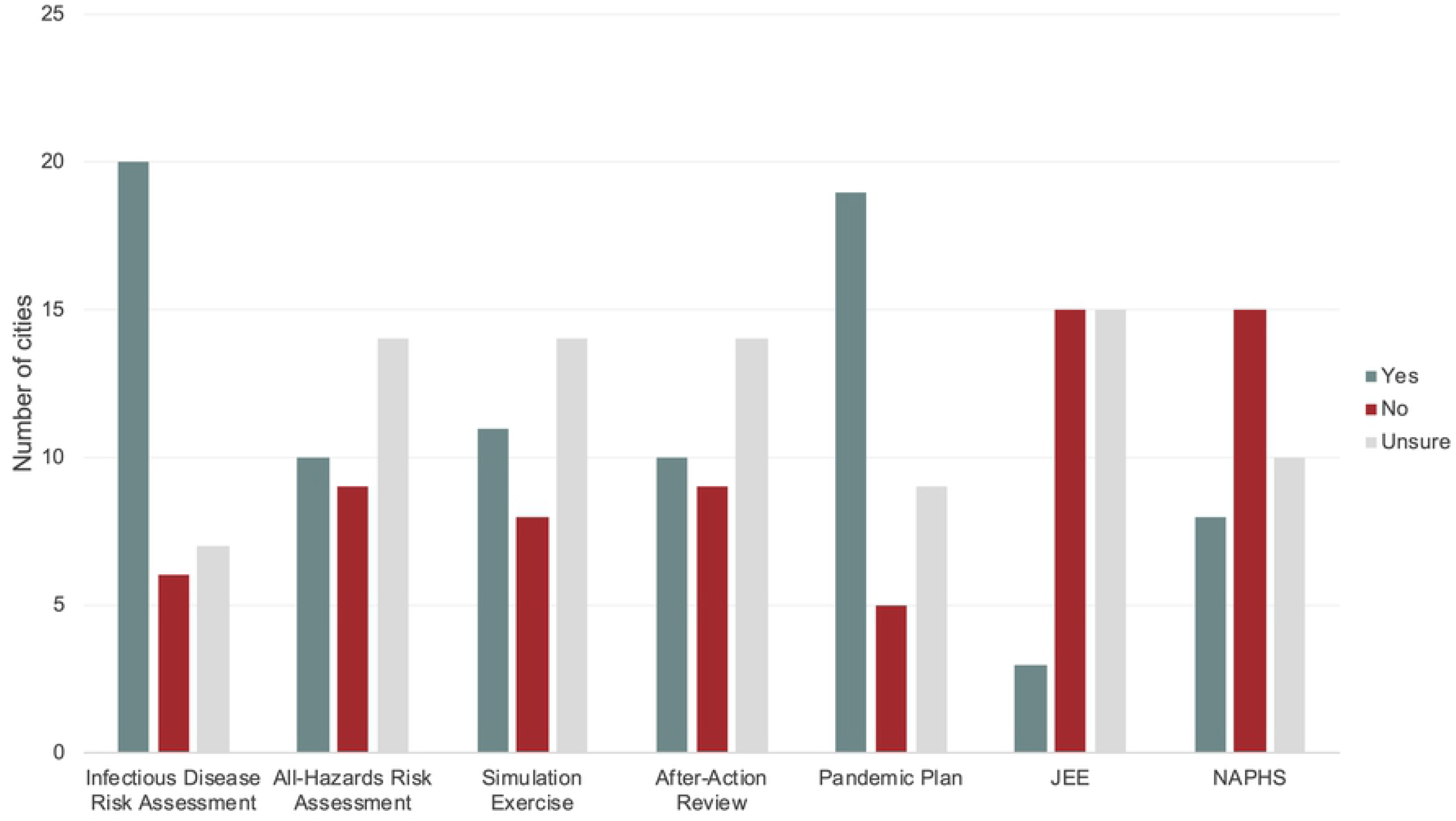
The number of cities that had completed or engaged in pandemic preparedness efforts prior to the COVID-19 pandemic; JEE – Joint External Evaluation of the International Health Regulations; NAPHS – National Action Plan for Health Security.

Three cities – Abidjan, Accra, Kumasi – reported that they had been involved in their country’s JEE, while eight cities – Abidjan, Accra, Addis Ababa, Amman, Bandung, Colombo, Kigali, and Kumasi – reported that they were included in the development of their countries’ respective National Action Plan for Health Security. Survey results are available for review in the S2 Appendix.

### Coordination of Epidemic and Pandemic Activities

Responses to the RACI matrices revealed a heterogeneous landscape of authorities, responsibilities, and communication networks. Some cities, such as Abidjan, Bangkok, and Montevideo reported that the national government maintained primary authority and was accountable for virtually all pandemic preparedness activities in the city; other cities, such as Bengaluru, Buenos Aires, and Vancouver reported that subnational governments were primarily accountable for pandemic preparedness activities; and other cities, such as Bandung, Chicago, and Kampala, reported that the local government was primarily accountable for pandemic preparedness activities. However, most often, the accountability and responsibility for pandemic preparedness activities were shared across the various levels of government. Reported RACI matrices are available for review in the S3 Appendix.

The activities national authorities were most often responsible for include developing pandemic preparedness and response policy (n=32), financing routine public health activities (n=28), and identifying and mapping resources required for the response to public health emergencies (n=25) (Fig 2); the activities subnational authorities were most often responsible for include operationalizing pandemic preparedness and response policy (n=28), developing and maintaining pandemic preparedness and response plans (n=27), and developing mechanisms for coordination between levels of government (n=26); the activities local authorities were most often responsible for include operationalizing pandemic preparedness and response policy (n=28), developing mechanisms for coordination between levels of government (n=25), and developing and maintaining pandemic preparedness and response plans (n=25). Relative to other the other levels of government, national authorities were most frequently reported to be responsible for three of the 11 pandemic preparedness activities (i.e., developing pandemic preparedness and response policy financing routine public health activities, and identifying and mapping resources required for the response to public health emergencies), subnational authorities for six of the 11 activities, and subnational authorities and local authorities for two of the 11 activities.

**Fig 2.**
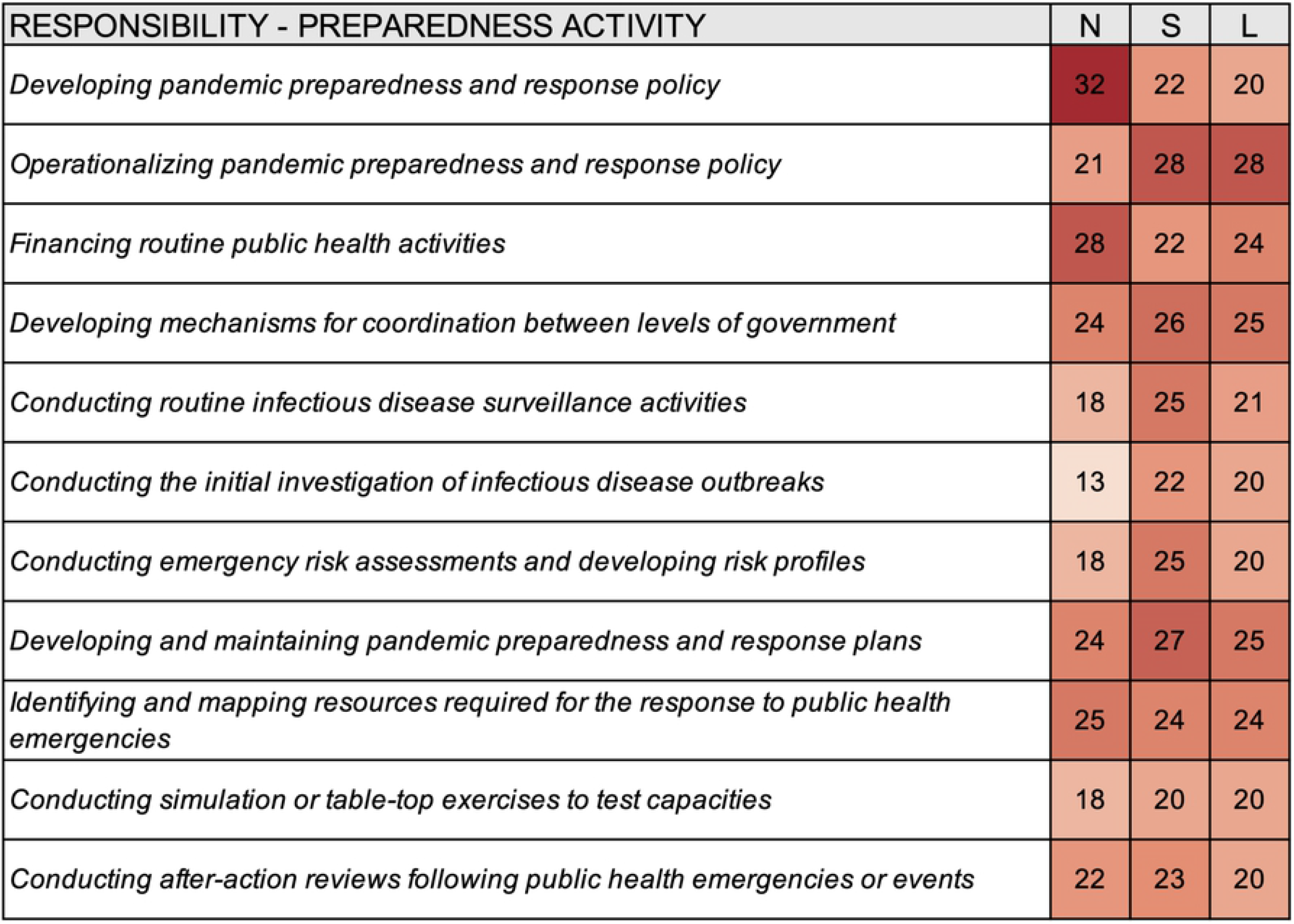
Heat map summarizing the responsibility for select preparedness activities for different levels of government according to survey responses from thirty-three cities; N – National, S – Subnational, L – Local.

Relative to the other levels of government, national authorities were reported to most often accountable for seven of the 11 pandemic preparedness activities, national and subnational authorities for one of the activities, subnational and local authorities for one of the activities, and local-level authorities for two of the activities (Fig 3). Yangon reported that no level of government was accountable for financing routine public health activities, conducting routine infectious disease surveillance activities, developing and maintaining pandemic preparedness and response plans, identifying and mapping resources required for the response to public health emergencies, and conducting simulation or table-top exercises to test capacities. Similarly, Athens reported that no level of government was accountable for conducting simulation or table-top exercises to test capacities and conducting after-action reviews following public health emergencies or events.

**Fig 3.**
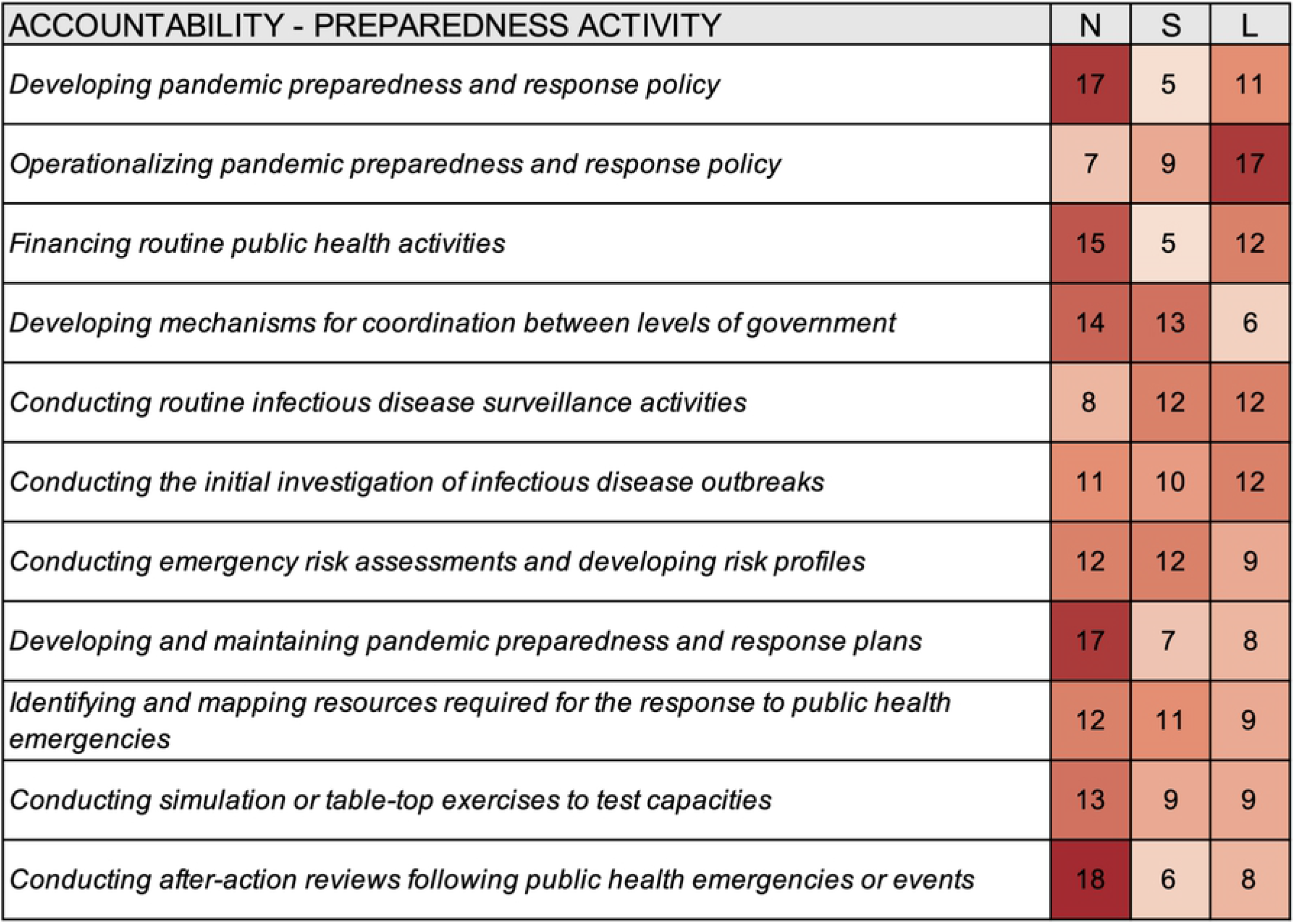
Heat map summarizing the accountability for select preparedness activities for different levels of government according to survey responses from thirty-three cities; N – National, S – Subnational, L – Local.

Survey responses indicated a relatively high degree of consultation for pandemic preparedness activities. National authorities were most often engaged in consultation for operationalizing pandemic preparedness and response policy (n=20), developing pandemic preparedness and response policy (n=14), and developing mechanisms for coordination between levels of government (n=14) (Fig 4); subnational authorities were most often engaged in consultation for developing pandemic preparedness and response policy (n=24), operationalizing pandemic preparedness and response policy (n=19), developing and maintaining pandemic preparedness and response plans (n=19), and conducting after-action reviews following public health emergencies or events (n=19); local authorities most often engaged in consultation for developing mechanisms for coordination between levels of government (n=15), developing and maintaining pandemic preparedness and response plans (n=14), and identifying and mapping resources required for the response to public health emergencies (n=13).

**Fig 4.**
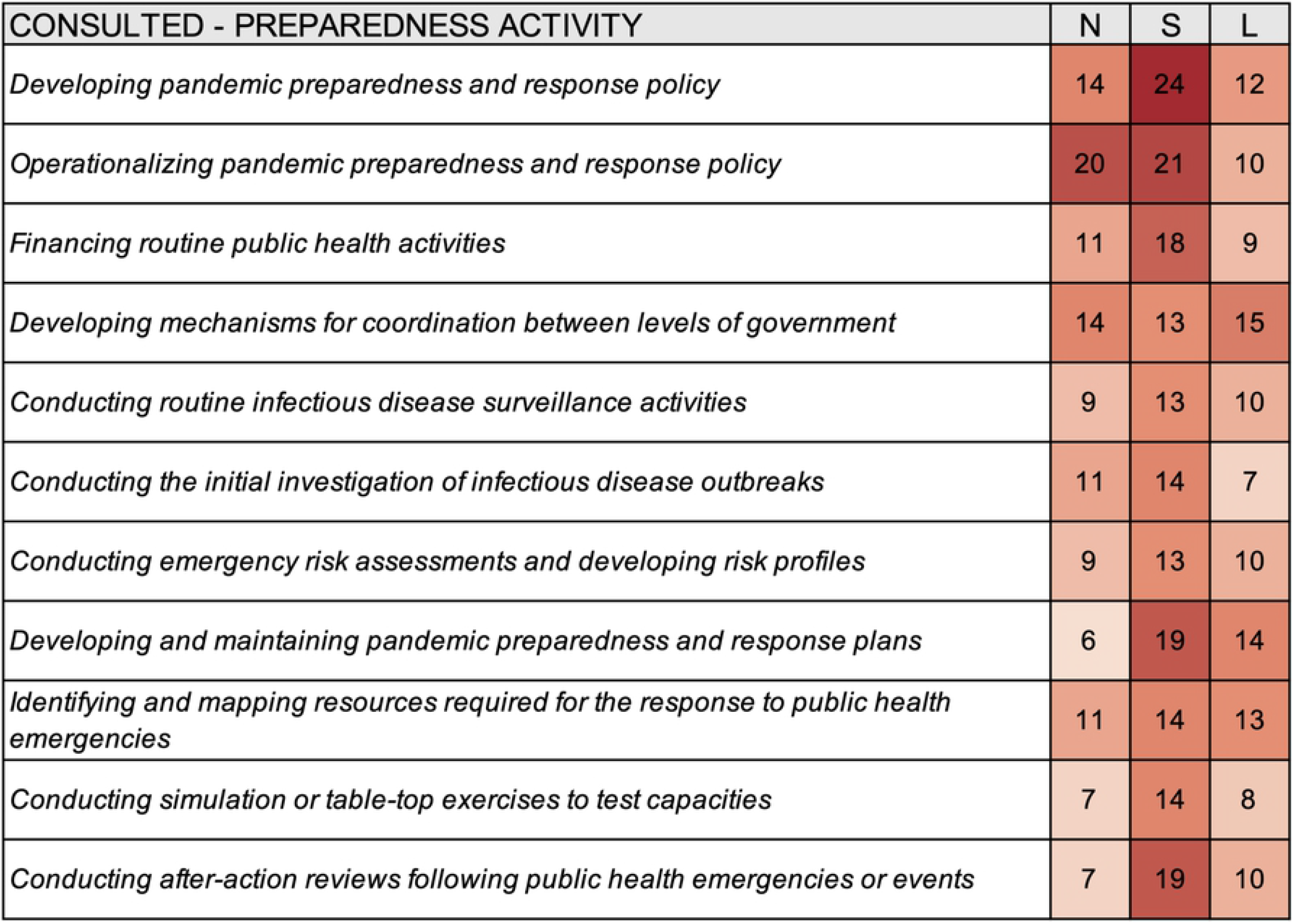
Heat map summarizing consultation for select preparedness activities for different levels of government according to survey responses from thirty-three cities; N – National, S – Subnational, L – Local.

Survey responses indicated a relatively low degree of one-way communication for preparedness activities. National authorities were most often informed regarding conducting routine infectious disease surveillance activities (n=6), conducting the initial investigation of infectious disease outbreaks (n=5), and conducting simulation or table-top exercises to test capacities (n=4) (Fig 5); subnational authorities were most often informed about conducting simulation or table-top exercises to test capacities (n=5), conducting after-action reviews following public health emergencies or events (n=4), financing routine public health activities (n=3), and conducting the initial investigation of infectious disease outbreaks (n=3); local authorities were most often informed regarding conducting simulation or table-top exercises to test capacities (n=4), financing routine public health activities (n=3), conducting the initial investigation of infectious disease outbreaks (n=3), conducting emergency risk assessments and developing risk profiles (n=3), and conducting after-action reviews following public health emergencies or events (n=3).

**Fig 5.**
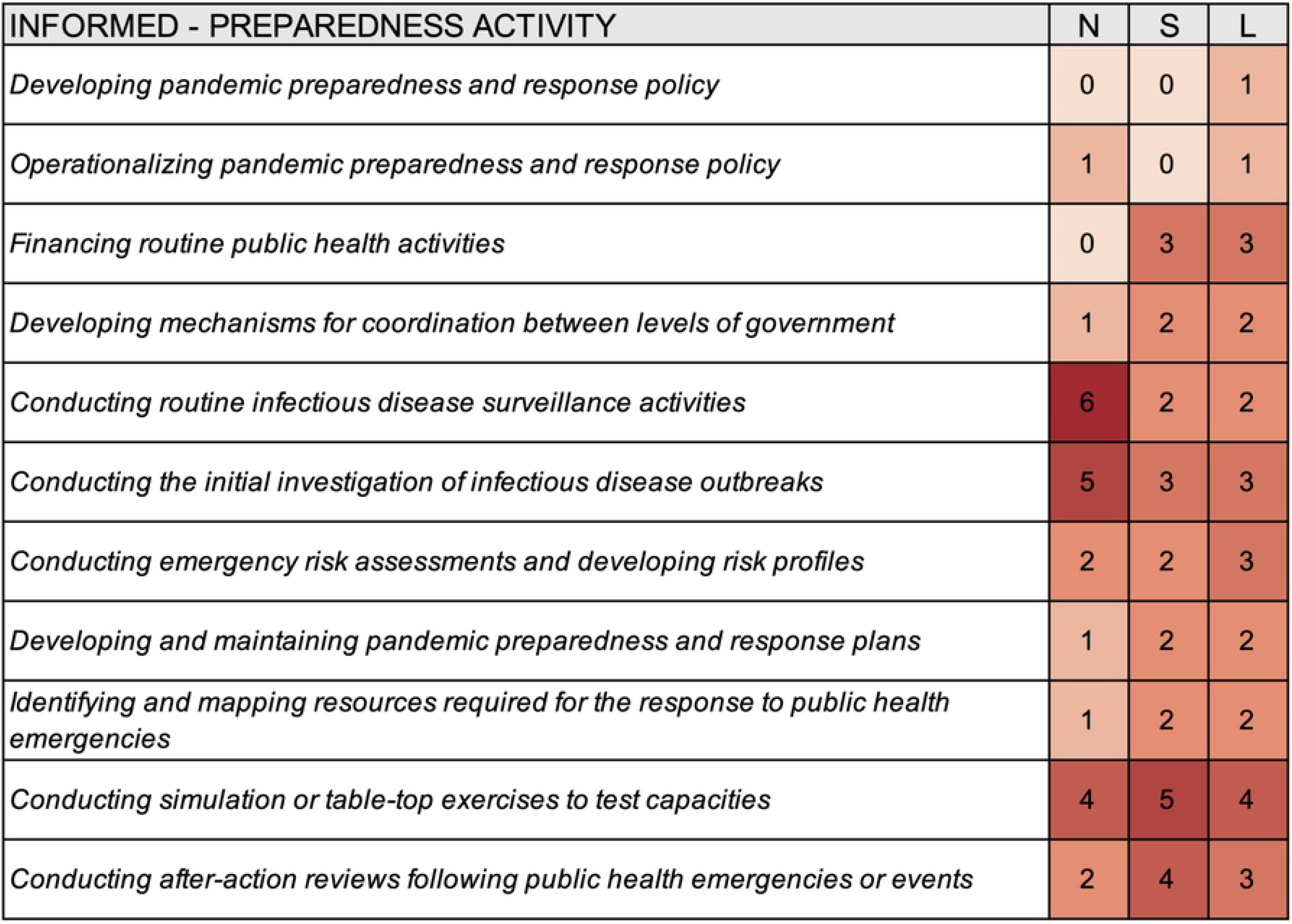
Heat map summarizing which levels of government are informed about select preparedness activities according to survey responses from thirty-three cities; N – National, S – Subnational, L – Local.

## Discussion

This study examined the ways in which local-level authorities were engaging in epidemic and pandemic preparedness efforts before the COVID-19 pandemic, as well as how they coordinated with higher levels of government for select preparedness activities. Answering these questions and better understanding the roles of local authorities in health security will be important as the world reflects on the experiences of COVID-19 and looks to prepare for future public health emergencies.

The results of this work highlight that, according to local governments and public health authorities, before the COVID-19 pandemic, epidemic and pandemic preparedness in cities was largely left to higher levels of government. Of the activities included in our survey, national governments were both responsible for implementing and accountable for overseeing a majority of the activities, when compared to subnational and local governments. Still, the role of local governments was not insignificant, as they were the level of government most often accountable for overseeing and responsible for operationalizing pandemic preparedness and response policy, and for overseeing routine infectious disease surveillance activities and conducting the initial investigation of outbreaks. They were also frequently consulted by higher levels of government as they developed mechanisms for coordination between levels of government, developed and maintained pandemic preparedness and response plans, and identified and mapped resources required for the response to public health emergencies.

These results corroborate the widely held view that local governments are often at the frontline of epidemic preparedness efforts – especially as they relate to infectious disease surveillance activities [25]. They also highlight that there are opportunities to better integrate local authorities into epidemic and pandemic preparedness efforts. While the specifics of how a health crisis is managed and who is involved will vary between cities and according to the governance context [20], at least 20 of the 33 participating cities reported that they were responsible for each of the preparedness activities considered in this study. The World Health Organization has implicitly acknowledged this responsibility and the vital role local governments play through their development of interim guidance for local authorities for strengthening preparedness for COVID-19 in urban environments [26].

Still, others have argued that the future success of global health interventions, may depend on the ability of local governments, especially those in cities, to promote health [18]. To this end, ensuring that cities are better prepared for future public health emergencies will require a renewed emphasis on strengthening local capacities [3]. Results from this study demonstrate that while local authorities do engage in proactive planning and risk assessment, they do not frequently participate in other efforts that are meant to measure preparedness or the functionality of existing capacities. This is, perhaps, unsurprising given that the current iterations of the assessment tools do not provide specific measures or guidance for subnational or local governments [27], and the view that national-level actors are primarily responsible for epidemic and pandemic response, which may include assessing the functionality of capacities.

To this end, should the COVID-19 pandemic prove impetus enough to reexamine how the world approaches measuring public health preparedness, one practical and impactful opportunity that exists for better including local governments in preparedness efforts would be to formally or informally incorporate them into existing preparedness assessment activities and tools. Alternatively, local governments could take it upon themselves to conduct preparedness assessments by using resources specifically designed for them [23], or by conducting voluntary local reviews – similar to how some have done for the Sustainable Development Goals [28]. These results, in addition to being an asset to local authorities, could also be used to help inform efforts at higher governance levels and provide context that is currently lacking [10,11].

Also of importance is that local governments, while frequently responsible or accountable for many of the preparedness activities, are not as frequently consulted or informed. This suggests that there are opportunities to improve governance, planning processes, financing, collaborative networks, and communication – all of which are essential elements of public health emergency preparedness [29]. Previous research investigating the effects of national-level government effectiveness, a key dimension of good governance, found that greater government effectiveness was associated with lower COVID-19 mortality rates [30]. Following the COVID-19 pandemic, national and subnational governments may wish to examine the ways in which they collaborate with local governments to complete key preparedness activities as a means of improving governance and effectiveness. These mechanisms for collaboration and communication should also be tested using robust simulation exercises that could help identify weaknesses, ambiguities, and bottlenecks [31].

This study and its results suffer from several limitations. First, and most significantly, the results relied on the completion of a survey by one individual. While we attempted to distribute the survey to the local authority who would be most knowledgeable about epidemic and pandemic preparedness in the city, we cannot guarantee that the data captured by responses are completely valid or factual. One person’s knowledge regarding all of the preparedness efforts, activities, and arrangements in their city may be limited. For instance, a majority of survey participants had served in their current professional role for one to four years and may not have been aware of preparedness efforts in their city before this time. Future research may wish to more deeply examine preparedness efforts in specific cities or to validate the results of this study by reviewing the legislation, regulations, and other legal frameworks that provide the foundation for public health preparedness and coordination between levels of government.

Additionally, the specific results from each city are unlikely to hold great amounts of external validity. The governance contexts in which participating cities exist vary widely, and it would be inappropriate to generalize results across contexts without examining the specific authorities of local-level authorities. Indeed, the powers of local governments can differ significantly between cities within the same country, let alone between cities in different countries. It is for this reason that our study provides descriptive statistics to summarize broad trends in urban health security in cities around the world, instead of attempting to overstate our results and risk inappropriately generalizing across contexts.

In conclusion, the results of this research suggest that, according to local-level authorities, their involvement in epidemic and public health preparedness before the COVID-9 pandemic was relatively limited. While many had completed risk assessments and developed some form of response plan, fewer had been involved in capacity assessments or tested the functionality of existing capacities through simulation exercises or after-action reviews. Further, while national governments seemingly represent the level of government that is primarily responsible and accountable for preparedness, local governments still maintained much responsibility and a large role in operationalizing policy and implementing activities. Opportunities for improving public health preparedness in the future may include improving coordination between levels of government, involving local-level authorities in capacity assessments, and examining the functionality of capacities at the local-level.

## Data Availability

All data used and/or analyzed during the current study are included in this published article and supporting information.

## Acknowledgements

Our thanks to Mariana Espinosa Estrada, Charity Hung, Ramya Kancharla, Sherissa Ng, Joseph Ngamije, Grace Pickens, and Tara O’Rourke of Vital Strategies for their assistance in the distribution of survey and study materials. Thanks also to Rebecca Katz of the Georgetown Center for Global Health Science & Security for her review of early drafts and comments that helped to improve this manuscript.

